# Incidence and outcomes of high-output heart failure in patients with arteriovenous fistula

**DOI:** 10.1101/2025.08.26.25334503

**Authors:** Alok Tripathi, Brandon Hanten, Muhammad Shafiq, Ankita Tiwari, Archana Gautam, Pratik Bhyan, Tarun Dalia, Bhanu Gupta

## Abstract

**Background:** Arteriovenous fistula (AVF) in patients with end-stage renal disease (ESRD) can lead to high-output heart failure (HOHF). There is limited data on the incidence and outcomes of HOHF in patients with AVF.

**Objective:** The main goal of our study was to determine the incidence and prevalence of HOHF [diagnosed via right heart catheterization (RHC)] in chronic kidney disease (CKD)/ESRD patients with AVF. We also aimed to evaluate the clinical determinants of the development of HOHF in this group.

**Methods:** We conducted a retrospective cohort study at the University of Kansas Medical Center from 2011 to 2023. Patients with CKD/ ESRD with AVF who underwent RHC after AVF creation were included in the study. HOHF was defined as a cardiac index (CI) ≥4.0 L/min/m², measured either with the Fick or the Thermodilution method. Bivariate and multivariable regression analyses were performed to identify independent predictors of HOHF in this population.

**Results:** Out of 84 patients with AVF, 34 patients met established inclusion and exclusion criteria. Ten out of 34 patients (29.4%) developed HOHF. Hemoglobin (Hb) was significantly lower in the HOHF group than the non-HOHF group (10.16 vs. 11.52 g/dL; p = 0.02). Patients with HOHF had significantly elevated CI when compared with the non-HOHF group (CI Fick: 4.54 vs. 2.91 L/min/m², p < 0.001). Similar mortality was observed in the HOHF and non-HOHF groups. After multivariant regression analysis, Hb was an independent predictor of HOHF (HR 0.86, 95% CI: 0.76-0.98, p=<0.01).

**Conclusion:** HOHF is common in patients with AVF; nearly 1/3^rd^ (29.4%) of the patients developed HOHF, confirmed with RHC. Low Hb was found to be an independent predictor of HOHF. Further larger studies are needed to confirm these findings and establish an early detection protocol to detect and treat this condition.

## INTRODUCTION

End-stage renal disease (ESRD) affects more than 600,000 people in the USA.^1^ An estimated 42% of the patients with functioning arteriovenous shunting develop symptoms of heart failure.^2^ Arteriovenous fistula (AVF) remains the gold standard for vascular access in patients ESRD, owing to its superior patency, reduced infection rates, and improved outcomes compared to central venous catheters and arteriovenous grafts.^1, 3, 4^ However, AVF can lead to a hemodynamically significant left-to-right shunt, characterized by a blood flow rate of 1 to 2 L/min, resulting in increased venous return, elevated cardiac preload, reduced systemic vascular resistance, and an overall increase in cardiac output (CO). These changes in hemodynamics can be pathophysiologically significant.^5, 6^ Although many patients adapt to these changes, a subset of ESRD/chronic kidney disease (CKD) patients, especially those with obesity, sleep disorder breathing, and lung disease, develop high-output heart failure (HOHF), a clinical condition where increased cardiac output fails to meet metabolic demands due to circulatory dysregulation or pathological remodeling.^5, 6^

The prevalence of HOHF in patients with AVFs in the study by *Saleh et al.* was as high as 24%. However, the diagnoses were based on echocardiographic criteria rather than invasive hemodynamics.^7^ Small observational studies have shown that patients with AVF have improved symptoms of pulmonary hypertension and heart failure after fistula ligation procedures, highlighting the clinical significance of this condition and its potential to become reversible with proper intervention.^8, 9^ Despite these findings, the literature is limited by small sample sizes, inconsistent definitions, and underuse of right heart catheterization (RHC), which is the gold standard for assessing cardiac output.^10^

Therefore, to better understand the burden and outcomes of AVF-associated HOHF, we conducted a retrospective cohort study at our center. Our research distinguishes itself by focusing on patients with RHC-confirmed HOHF. The primary objective of our study was to determine the frequency of HOHF in patients with CKD/ESRD after AVF creation. Additionally, the study aimed to accomplish two additional objectives: to identify risk factors for developing HOHF through clinical and laboratory assessments, and to evaluate one-year mortality and follow-up outcomes.

## MATERIALS AND METHODS

### Study design

This study was conducted at the University of Kansas Medical Center, Kansas City, Kansas. The study used a retrospective cohort design. First, using the inclusion criteria outlined below, database queries were conducted for patients observed in the health system between January 2011 and December 2023 using the Healthcare Enterprise Repository for Ontological Narration (HERON). This search discovery tool enables cohort building for observational research. Once the initial cohort was identified using the inclusion criteria, the medical record numbers for those patients were obtained with the help of the Clinical Informatics department. Subsequently, individual patient charts were reviewed for data collection. Institutional Review Board (IRB) approval was required and was obtained from the study site (IRB approval number# STUDY00148490)

### Selection criteria

#### Inclusion criteria

1. CKD/ ESRD with AVF with a documented history of heart failure based on the ICD diagnosis.
2. Patients who had RHC done after AV fistula creation.

#### Exclusion criteria

1. Age>85 years or <18 years.
2. Patients with other causes of HOHF, like liver cirrhosis/ thyrotoxicosis at the time of RHC, anemia with Hemoglobin <8 g/dL at the time of RHC, or obesity with BMI >35 kg/m2.
3. Patients who had an iatrogenic fistula as a complication of a medical procedure (like Cardiac Cath) involving vascular access.
4. Patients with a spontaneous/congenital fistula.

### High-output heart failure

A cardiac index of 4.0 L/min/m² or higher defined HOHF when measured by either Fick or Thermodilution method.

### Outcome

This study evaluated the incidence of HOHF in patients who underwent arteriovenous fistula implantation for hemodialysis or lipid apheresis treatment.

#### Statistical analysis

##### Bivariate analysis

This research examined the relationship between HOHF and the following variables, in addition to essential demographic factors such as age, sex, and race:

- Time interval from arteriovenous fistula (AVF) creation to RHC, measured in months
- Presence of coronary artery disease
- Presence of atrial fibrillation
- Personal history of hypertension
- History of diabetes mellitus
- Use of beta-blockers
- Use of ACE-Inhibitor (ACE-I) or Angiotensin receptor blocker (ARB)
- Use of diuretics
- Hemoglobin levels were recorded based on the value reported closest to the time of right heart catheterization (RHC).
- Thyroid-stimulating hormone (TSH) levels were obtained from the measurement closest to the timing of RHC.
- Left ventricular ejection fraction (LVEF) was assessed using echocardiographic data available nearest to the time of RHC.
- Follow-up interval after RHC
- Mortality data

The association between HOHF and categorical variables (including sex, race, coronary artery disease, atrial fibrillation, hypertension, diabetes, use of beta-blockers, use of ACE-I or ARB, and use of diuretics) was evaluated using Fisher’s exact test.

The age difference, along with hemoglobin and HOHF, was analyzed using *a t*-test. The time from fistula creation to the RHC (in months) did not follow a normal distribution. Similarly, the TSH level and LVEF also did not follow a normal distribution. Therefore, the association between HOHF and the time from fistula creation to the RHC, TSH level, and LVEF was assessed using the Wilcoxon signed-rank test. The TSH level was missing for two patients, and there was one outlier in the TSH level. These three patients (two with missing data and one with an outlier TSH level) were excluded only from the comparison between TSH levels and HOHF. The alpha criterion was set at 0.05 for all statistical tests.

##### Regression analysis

Parametric binomial regression was initially attempted, but it was noted that many continuous variables did not have a normal distribution (as discussed above). Even when those variables (which did not have a normal distribution) were removed from the model, parametric binomial regression did not converge, likely due to the small sample size and the large number of independent variables. Therefore, an alternative approach, non-parametric binomial regression using bootstrap with quantile regression, was employed. Except for race (due to very small count in more than one subgroup), hypertension (as all patients in HOHF group had hypertension) and TSH level (due to missing data), all other variables (including age, sex, time from fistula creation to right-side heart catheterization, coronary artery disease, atrial fibrillation, diabetes mellitus, use of beta-blockers, use of diuretics, hemoglobin level, TSH level and LVEF) were used as the independent variables in the final regression model.

## RESULTS

A total of 84 patients with AV fistulas were initially identified based on the inclusion criteria. Of these 84 patients, 50 were excluded according to the previously mentioned criteria. The final cohort consisted of 34 patients. Among the final 34 patients, HOHF was observed in 10 patients (29.41%). The basic characteristics of the final cohort are detailed in **Table 1**.

**Table 1:**
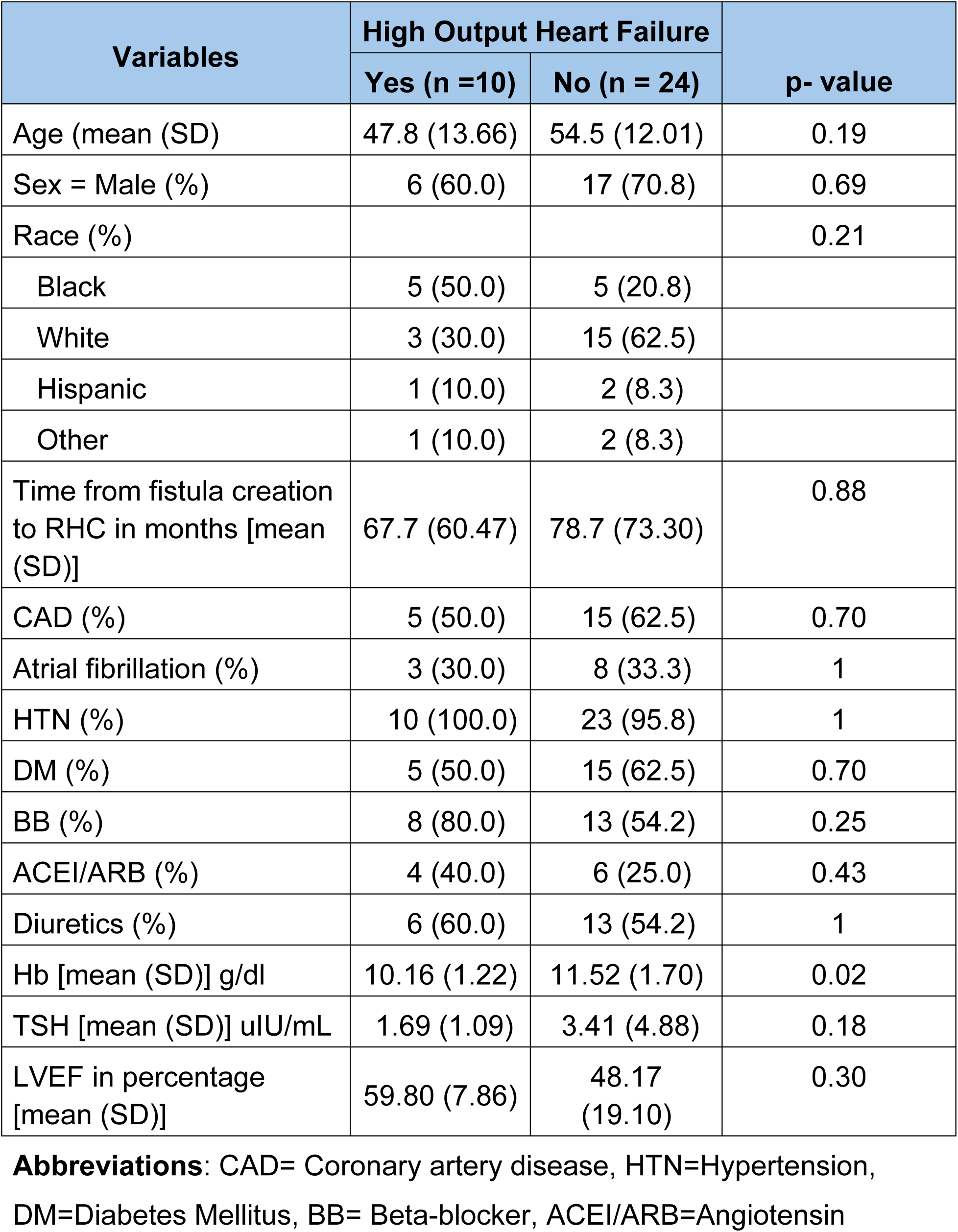
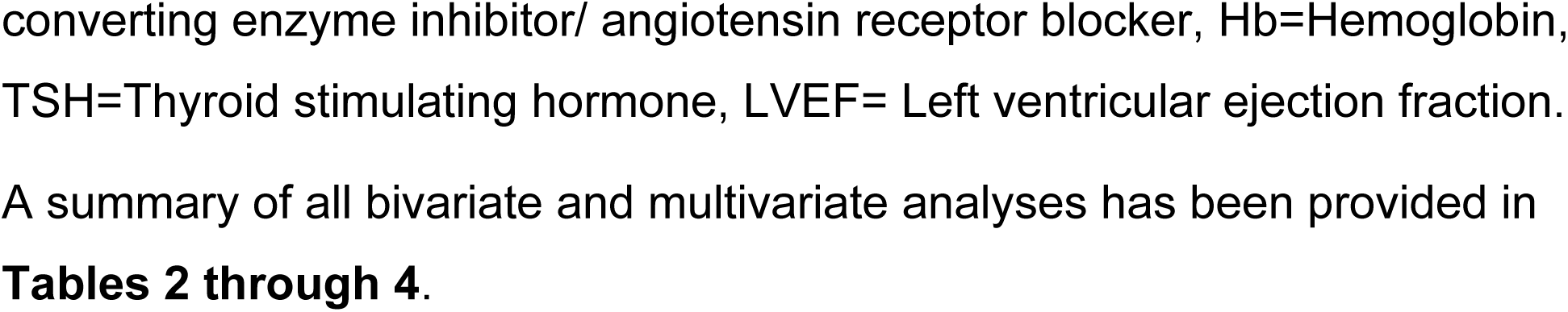
Basic characteristics of the final cohort.

The differences in hemodynamic profiles between the two groups are detailed in Table 6. A significant distinction was the higher CO and CI values observed with both the Fick and thermodilution methods. Using the Fick method, the HOHF group versus the no-HOHF group showed higher CO (7.93 vs 5.96, p <0.01) and higher CI (4.54 vs 2.91, p <0.01). Similarly, the HOHF group had higher CO (7.30 vs 5.36, p <0.01) and higher CI (4.14 vs 2.61, p <0.01) according to the thermodilution method.

**Table 2:** Bivariate and Multivariant analysis of categorical variables to predict HOHF.

| Variables | Hazard Ratio | 95% CI for Risk Ratio |  | p-value |
| --- | --- | --- | --- | --- |
|  |  | Lower Limit | Upper Limit |  |
| **Bivariant Analysis** |  |  |  |  |
| Sex (Male) | 0.72 | 0.25 | 2.03 | 0.69 |
| Race |  |  |  | 0.21 |
| Black | Reference |  |  |  |
| White | 0.33 | 0.08 | 1.07 |  |
| Hispanic | 0.66 | 0.04 | 2.50 |  |
| Other | 0.66 | 0.04 | 2.50 |  |
| CAD | 0.70 | 0.25 | 1.97 | 0.70 |
| Atrial fibrillation | 0.90 | 0.28 | 2.82 | 1 |
| DM | 0.70 | 0.25 | 1.97 | 0.70 |
| BB | 2.48 | 0.62 | 9.91 | 0.25 |
| ACEI or ARB | 1.60 | 0.57 | 4.47 | 0.43 |
| Diuretics | 1.18 | 0.41 | 3.45 | 1 |
| **Multivariate**<br>**Analysis:** |  |  |  |  |
| Age (in years) | 0.98 | 0.96 | 1.00 | 0.02 |
| Sex (male) | 0.94 | 0.61 | 1.39 | 0.43 |
| CAD | 1.30 | 0.81 | 1.94 | 0.16 |
| Afib | 1.13 | 0.78 | 1.56 | 0.42 |
| DM | 1.06 | 0.73 | 1.42 | 0.42 |
| ACEi or ARB Use | 1.10 | 0.71 | 1.66 | 0.38 |
| Beta-Blocker Use | 1.00 | 0.64 | 1.59 | 0.46 |
| Diuretics Use | 1.22 | 0.79 | 1.93 | 0.25 |
| Time from Fistula<br>Creation to RHC (in<br>months) | 1.00 | 0.99 | 1.00 | 0.16 |
| Hemoglobin (g/dL) | 0.86 | 0.76 | 0.98 | **<0.01** |
| LVEF (%) | 1.01 | 1.00 | 1.02 | **<0.01** |

**Abbreviations:** CAD= Coronary artery disease, Afib= Atrial Fibrillation, DM=Diabetes Mellitus, ACEI/ARB=Angiotensin converting enzyme inhibitor/ angiotensin receptor blocker, LVEF= Left ventricular ejection fraction

**Table 3:** Bivariate analysis of continuous variables with normal distribution to predict HOHF.

| Variables | Means Difference | 95% CI for Means Difference |  | p-value |
| --- | --- | --- | --- | --- |
|  |  | Lower Limit | Upper Limit |  |
| Age (mean (SD)) | 6.74 | -3.84 | 17.32 | 0.19 |
| Hemoglobin (mean (SD)) | 1.36 | 0.28 | 2.43 | 0.02 |

**Table 4:**
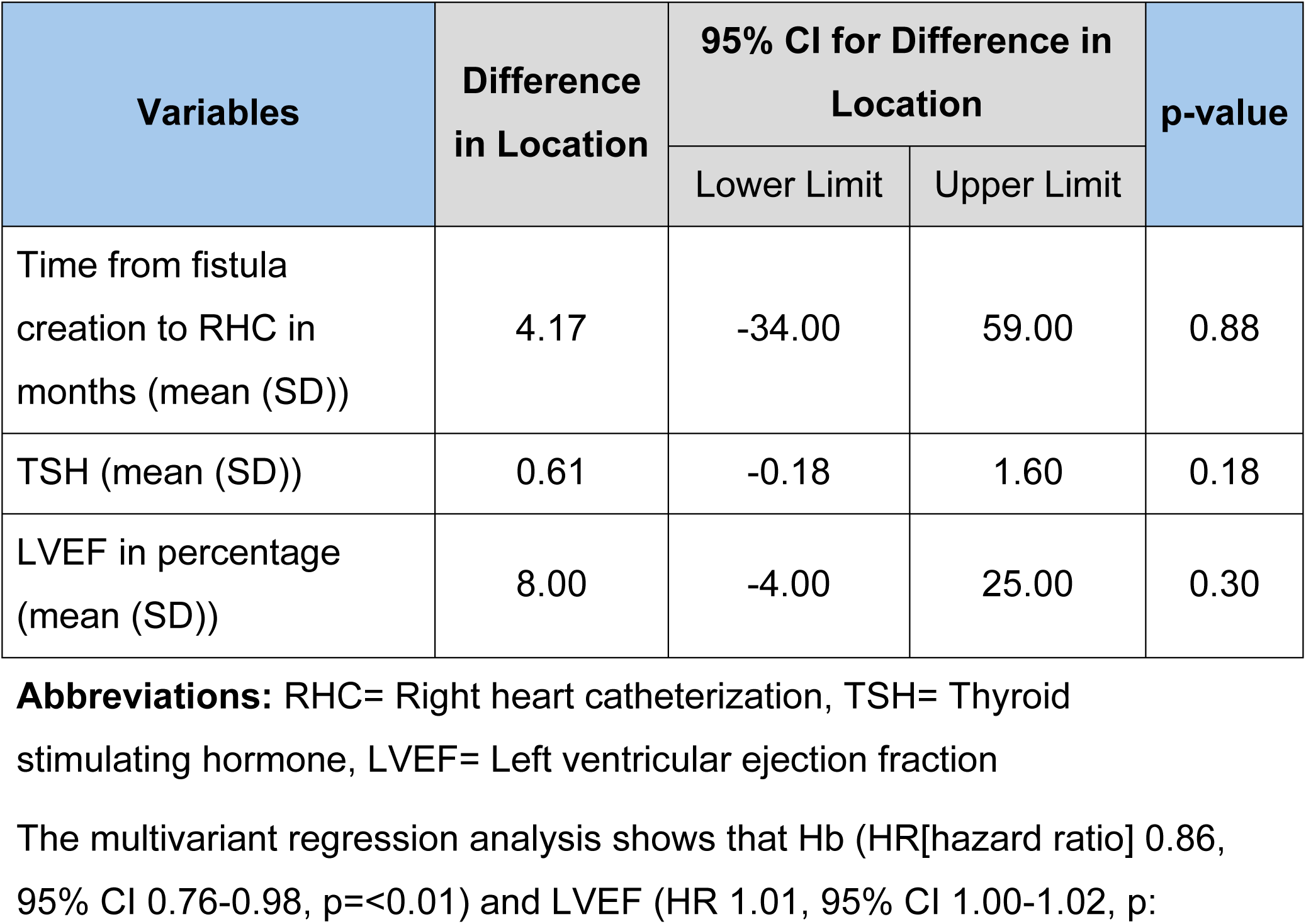
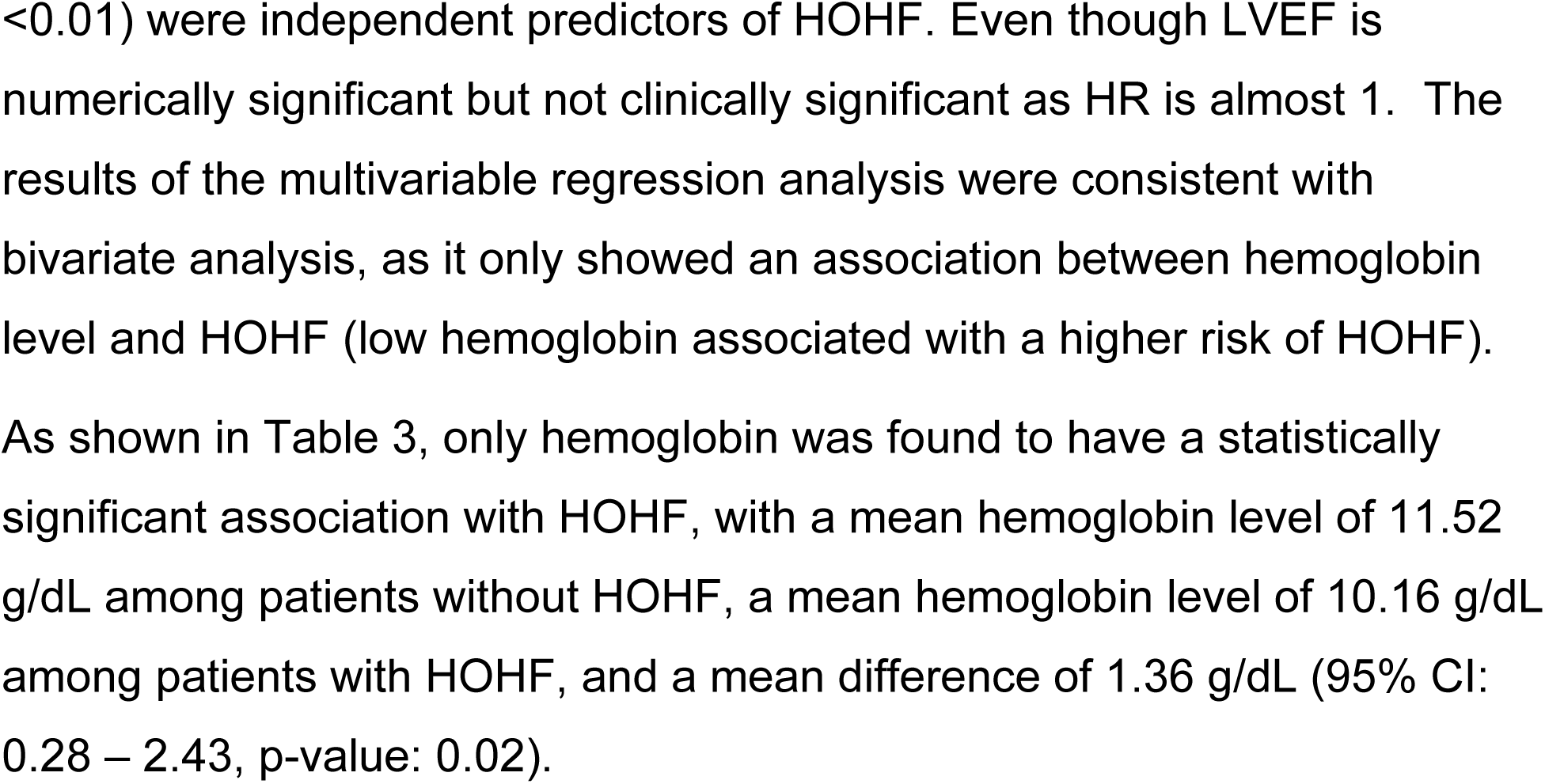
Bivariate analysis of continuous variables without normal distribution to predict HOHF.

**Table 5:**
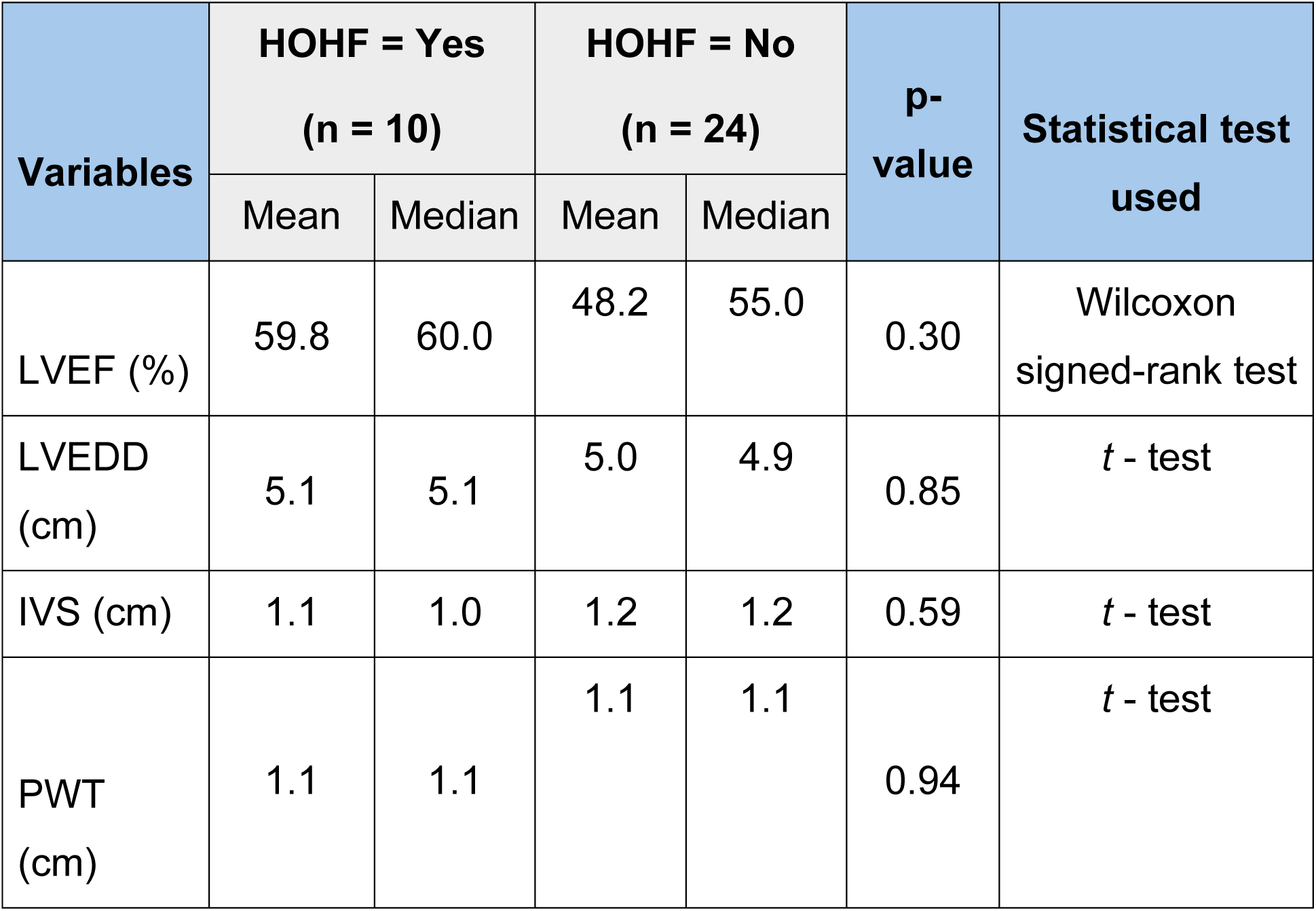
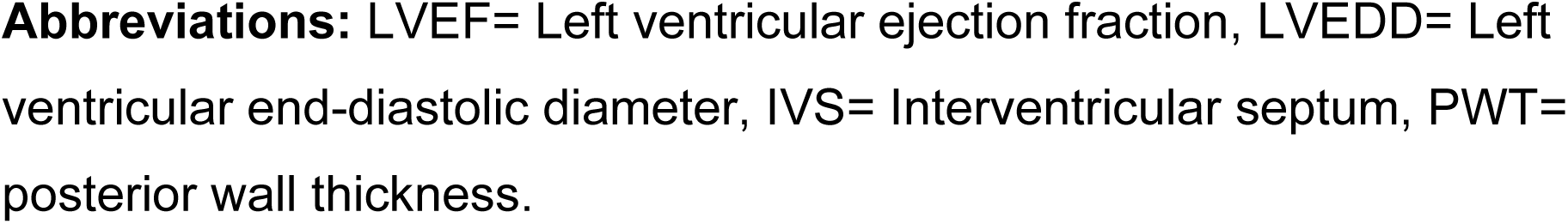
Echocardiogram data comparison between HOHF and no-HOHF group.

**Table 6:** RHC Data Comparison between patients with HOHF and without HOHF.

| RHC Data Elements | HOHF |  | p-value |
| --- | --- | --- | --- |
|  | Yes (n = 10) | No (n = 24) |  |
| RA (mean (SD)) mmHg | 11.40 (5.82) | 10.21 (4.93) | 0.55 |
| PA Mean (mean (SD)), mmHg | 33.90 (7.37) | 27.62 (9.11) | 0.06 |
| PCWP (mean (SD)) | 17.40 (6.96) | 16.50 (7.48) | 0.75 |
| CO Fick (mean (SD)) L/min | 7.93 (1.21) | 5.96 (1.82) | **<0.01** |
| CI Fick (mean (SD)) L/min/m<sup>2</sup> | 4.54 (0.89) | 2.91 (0.79) | **<0.01** |
| CO Thermo (mean (SD)) | 7.30 (1.16) | 5.36 (2.10) | **0.01** |
| CI Thermo (mean (SD)) | 4.14 (0.73) | 2.61 (0.93) | **<0.01** |
| PVR (mean (SD)) | 2.15 (0.81) | 2.07 (1.05) | 0.86 |

**Abbreviations:** RA= Right atrial, PA= Pulmonary artery, PCWP= Pulmonary capillary wedge pressure, CO= Cardiac output, CI= Cardiac index, PVR= Pulmonary vascular resistance.

### Mortality comparison

Among patients with HOHF, 30% (n=3) died in follow-up, and among patients without HOHF, 54% (n=13) died during follow-up. The risk ratio of mortality among patients with HOHF when compared to patients without HOHF is 0.55 (95% CI: 0.20, 1.53, p-value: 0.27, Fisher exact test)

### Follow-up comparison (in months)

The mean (SD) follow-up duration among patients with HOHF was 107 (76.22) months, while for patients without HOHF, it was 86.71 (82.94) months.

## DISCUSSION

Our study offers novel and clinically meaningful insights into the burden of HOHF among patients with ESRD after AVF creation. To our knowledge, this is the only study that has assessed HOHF in this specific population using invasive hemodynamic data from RHC. Key findings include: 1) The prevalence of HOHF remains high among patients with AV fistula (29.4%), 2) low hemoglobin levels were significantly associated with the development of HOHF in AV fistula patients.

The prevalence of HOHF was found to be 29.4% in our AV fistula patient group based on cardiac index measurements exceeding 4.0 L/min/m² obtained through RHC.^11^ Our study shows that patients with AV fistula have a high incidence of HOHF. These prevalence findings are similar to the 24% rate that *Saleh et al.* reported. However, those findings were based on echocardiographic criteria rather than direct invasive measurements by right heart catheterization (RHC).^7^ Additionally, *Reddy et al*., in an extensive 15-year single-center study evaluating HOHF through RHC in a broader patient population, identified that 22% of HOHF cases were attributable to arteriovenous shunts.^12^ The use of RHC in our study enhances diagnostic specificity compared to prior observational studies, many of which rely on echocardiographic surrogates or clinical judgment. RHC remains the gold standard for characterizing systemic vascular resistance and cardiac output and should be more routinely employed in symptomatic ESRD/CKD patients with AVFs to diagnose HOHF.^13^

Among the clinical and laboratory variables assessed in this study, hemoglobin concentration emerged as the exclusive parameter with a statistically significant association with HOHF. Patients with HOHF had significantly lower average hemoglobin levels than patients without HOHF, with values of 10.16 g/dL and 11.52 g/dL, respectively (p = 0.02). The findings indicate that anemia plays a significant role in causing high-output states among patients with AVFs. Anemia lowers blood oxygen delivery by reducing its oxygen-carrying capacity; therefore, the body responds with increased cardiac output to maintain tissue oxygen supply.^14^ When this compensatory response is superimposed upon the already elevated preload and cardiac output associated with AVF-induced shunting, the risk of HOHF is further amplified. Although no studies to date have specifically reported an association between anemia and AVF-associated HOHF, prior research has shown that chronic anemia is associated with increased left ventricular thickness in ESRD patients.^15, 16^ In addition, multiple studies have demonstrated that anemia worsens heart failure with preserved ejection fraction (HFpEF), contributing to increased mortality and hospitalizations.^17, 18^ Taken together, these findings suggest that anemia is not merely a comorbidity but may act as a modifiable risk factor in the pathogenesis of AVF-associated HOHF.

In our retrospective analysis, common comorbidities and clinical characteristics, including coronary artery disease, atrial fibrillation, hypertension, diabetes mellitus, medication use, and time since AVF creation, did not significantly differ between patients with and without HOHF. This lack of statistical association may result from the study’s limited sample size or could indicate a different pathophysiological mechanism in these patient populations, where anemia and volume overload potentially exert a more dominant influence than pre-existing structural heart disease. Interestingly, 100% the HOHF patients carried a diagnosis of hypertension, which demonstrates the complex nature of vascular resistance together with fluid status and cardiac function in patients with CKD/ESRD. The hemodynamic data showed that patients in the HOHF group presented characteristics of a hyperdynamic condition.

Patients with HOHF showed significantly higher cardiac output (CO) and cardiac index (CI) measurements, as determined by both the Fick and thermodilution methods. The patients with HOHF had a mean CI Fick value of 4.54 L/min/m², compared to 2.91 L/min/m² in those without HOHF (p < 0.001). HOHF patients also exhibited slightly higher mean pulmonary artery (PA) pressures (33.9 mmHg vs. 27.6 mmHg), although the difference was not significant due to the small sample size. Pulmonary hypertension (PH) in ESRD patients is a clinically significant finding, as it has been independently linked to increased mortality in this population.^19^

Cardiac output increases proportionally with the size of the AVF, and larger AVFs are linked to a higher risk of HOHF.^20^ This increased flow through AVF causes ongoing sympathetic activation, a higher heart rate, and myocardial contractility.^21^ Chronic overload induces cardiac remodeling, characterized by chamber dilation and the development of eccentric left ventricular hypertrophy (LVH).^5, 22^ LVH progression is linked to cardiovascular (CV) morbidity, including myocardial fibrosis and diastolic dysfunction, CV events, and all-cause mortality.^23, 24^ Interestingly, AVFs decrease the subendocardial viability index, thereby increasing myocardial oxygen demand while limiting coronary perfusion. This physiological imbalance puts hemodialysis patients at risk for subendocardial ischemia, myocardial stunning, and eventually, cardiomyopathy.^25, 26^

Although our study did not show a statistically significant difference in one-year mortality (risk ratio: 0.55; *p* = 0.27), this may be due to the small sample size of our study. Some earlier studies have reported poor long-term outcomes for patients with AVF-associated HOHF. Five-year mortality was reported at 58%, according to *Reddy et al*., in shunt-associated HOHF.^12^ In addition, Warner et al. recently published an expert opinion highlighting the clinical importance of timely diagnosis and intervention for AVF-induced HOHF to prevent cardiac remodeling and pulmonary hypertension progression.^5^

Our research confirms the emerging awareness about HOHF as an essential but frequently overlooked condition affecting patients with ESRD who have functioning AVFs. Given the superior diagnostic precision of RHC, invasive hemodynamic assessment should be considered in symptomatic CKD/ESRD patients with AVF, particularly when HOHF is suspected. While AVFs remain the preferred modality for long-term hemodialysis access, a more individualized approach is warranted, one that balances access durability with the potential for adverse cardiac remodeling. Larger prospective studies with serial echocardiography, appropriate utilization of RHC, and extended follow-ups are needed to establish standardized criteria for diagnosis, risk stratification, and ideal management of this underappreciated but clinically significant condition.

### Limitations

Several limitations must be acknowledged when interpreting the study’s results. The retrospective design introduces selection bias issues, which limit the ability to determine how AVF creation and anemia influence the development of HOHF. The detailed hemodynamic data RHC used in this study had limitations because they were only obtained from patients who required RHC for clinical reasons, potentially not representing the broader ESRD/CKD population with AVF.

The study had a small patient population because only 34 patients fulfilled all the criteria. The small sample size made it challenging to identify connections between HOHF and other variables. But the similar statistical analysis methods in small sample studies has been used in prior published studies. Some additional variables, such as AVF flow rates and BNP measurements, which could improve our understanding of HOHF pathophysiology, were unavailable during the study period and thus were not included in the analysis.

Lastly, the lack of standardized diagnostic criteria for HOHF across institutions limits the generalizability of our findings. Although we used a cardiac index threshold greater than 4.0 L/min/m² to define HOHF, this remains a topic of ongoing debate in the literature. Future prospective studies with standardized protocols and larger cohorts are necessary to validate these observations and clarify clinical thresholds for diagnosis and intervention.

## CONCLUSION

Our study results indicate that HOHF is common after AVF formation in patients with CKD/ESRD. The prevalence of HOHF among patients with AVF reached nearly 1 in 3 people, highlighting its importance for clinical assessment in patients with AVF and volume overload or heart failure symptoms. Anemia was independently and significantly associated with the development of HOHF. Hence, routine hemoglobin level monitoring and correction can be critical components for heart failure management in patients with CKD and ESRD.

## Data Availability

Data is restricted as per IRB insitution policy.

## BIBLIOGRAPHY

AVF: Arteriovenous fistula
ESRD: End-stage renal disease
CKD: Chronic kidney disease
HOHF: High-output heart failure
LVH: Left ventricular hypertrophy
CV: Cardiovascular
RHC: Right heart catheterization.
ACE-I: ACE-Inhibitor
ARB: Angiotensin receptor blocker
HFpEF: Heart failure with preserved ejection fraction.
PH: Pulmonary Hypertension
LVEF: Left ventricular ejection fraction

